## Supplemental Figures for "Divergent landscapes of A-to-I editing in postmortem and living human brain"

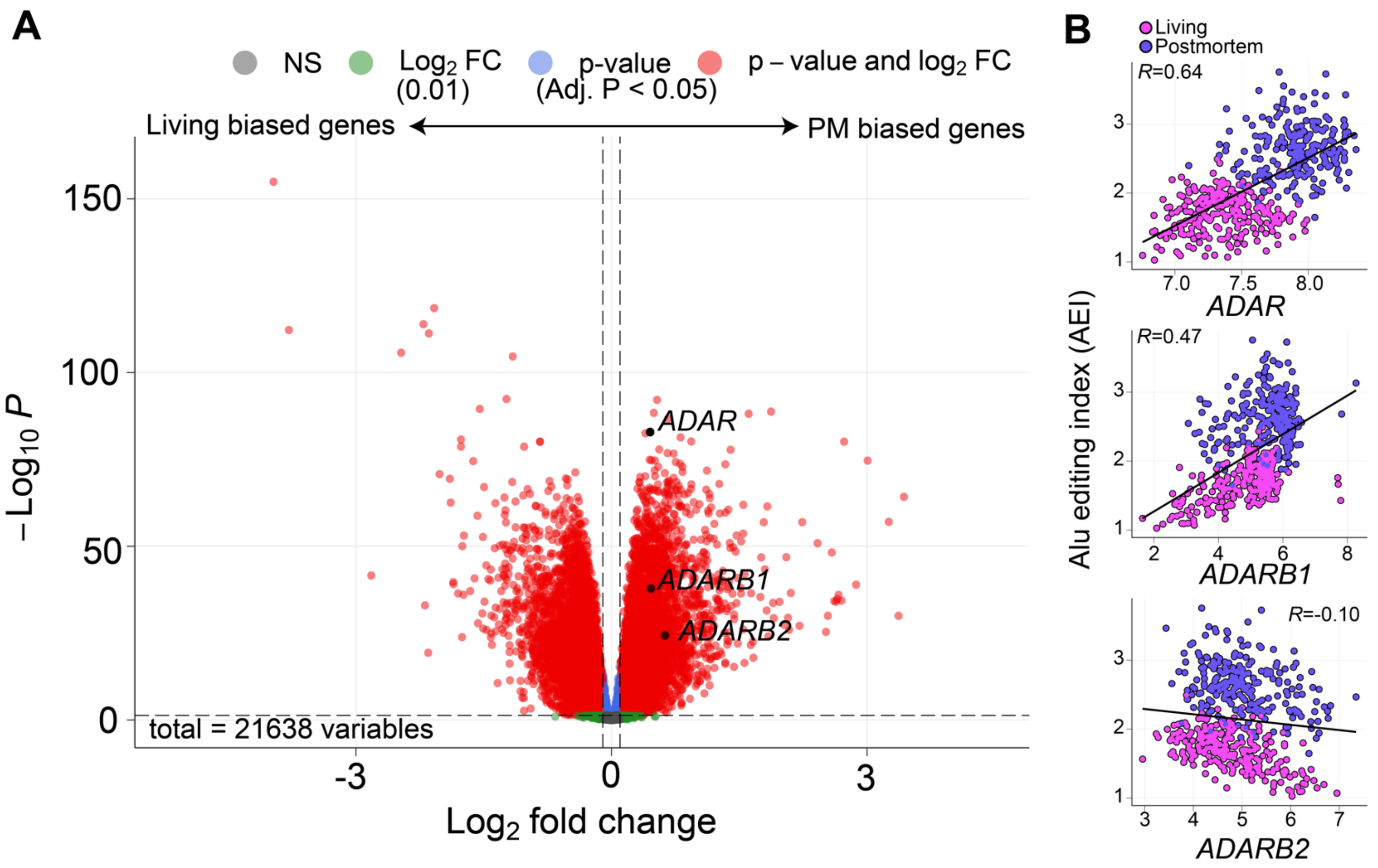


**Supplemental Figure 1. Transcriptome-wide differential gene expression between postmortem and living DLPFC.** (**A**) Volcano plot of differentially expressed genes between postmortem (PM) and living DLPFC compares $\log_2$ fold change (x-axis) with strength of significance ($-\log_{10}$ adjusted p-value, y-axis). Genes *ADAR*, *ADARB1* and *ADARB2* are colored black to visualize their effect size differences relative to the remaining transcriptome. (**B**) Pearson's correlation coefficients between the AEI and normalized gene expression (log2CPM) for *ADAR* (top), *ADARB1* (middle) and *ADARB2* (bottom).

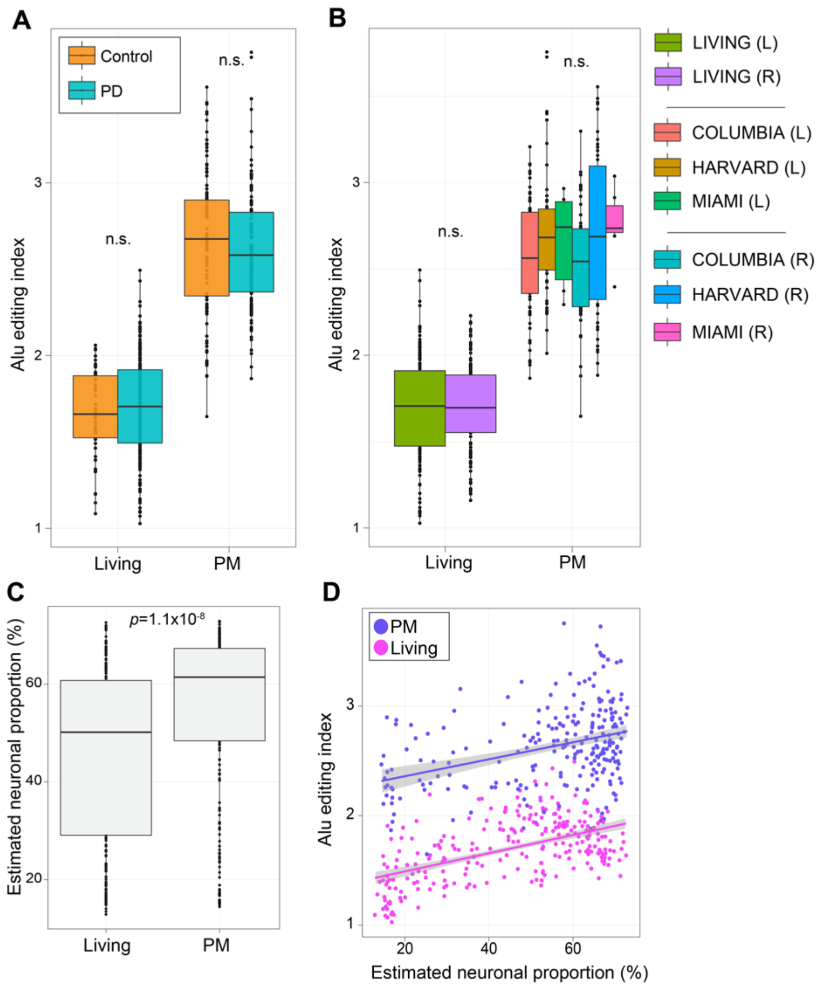


**Supplemental Figure 2. The effect of medical diagnosis, biobank and neuronal content on *Alu* editing.** (**A**) The *Alu* editing index (AEI; y-axis) between living and postmortem (PM) DLPFC parsed by individuals with Parkinson's disease (PD) and controls (x-axis). Two-sided linear regression was used to test for significance. No significant differences (n.s.) were observed. (**B**) The AEI (y-axis) between living and PM DLPFC parsed by different brain banks (x-axis). A Kruskal–Wallis test was used to test for significance among PM DLPFC samples. Further, differences between the left and the right hemisphere were tested within living DLPFC using a two-sided linear regression. No significant differences (n.s.) were observed. (**C**) dtangle estimated neuronal cell type proportions (y-axis) between living and PM DLPFC (x-axis). Two-sided linear regression was used to test for significance. All boxplots in this figure show the medians (horizontal lines), upper and lower quartiles (inner box edges), and 1.5× the interquartile range (whiskers). (**D**) Estimated neuronal proportions (y-axis) relate to changes in the AEI within both living and PM DLPFC (x-axis). A Pearson's correlation coefficient was used to test each association.

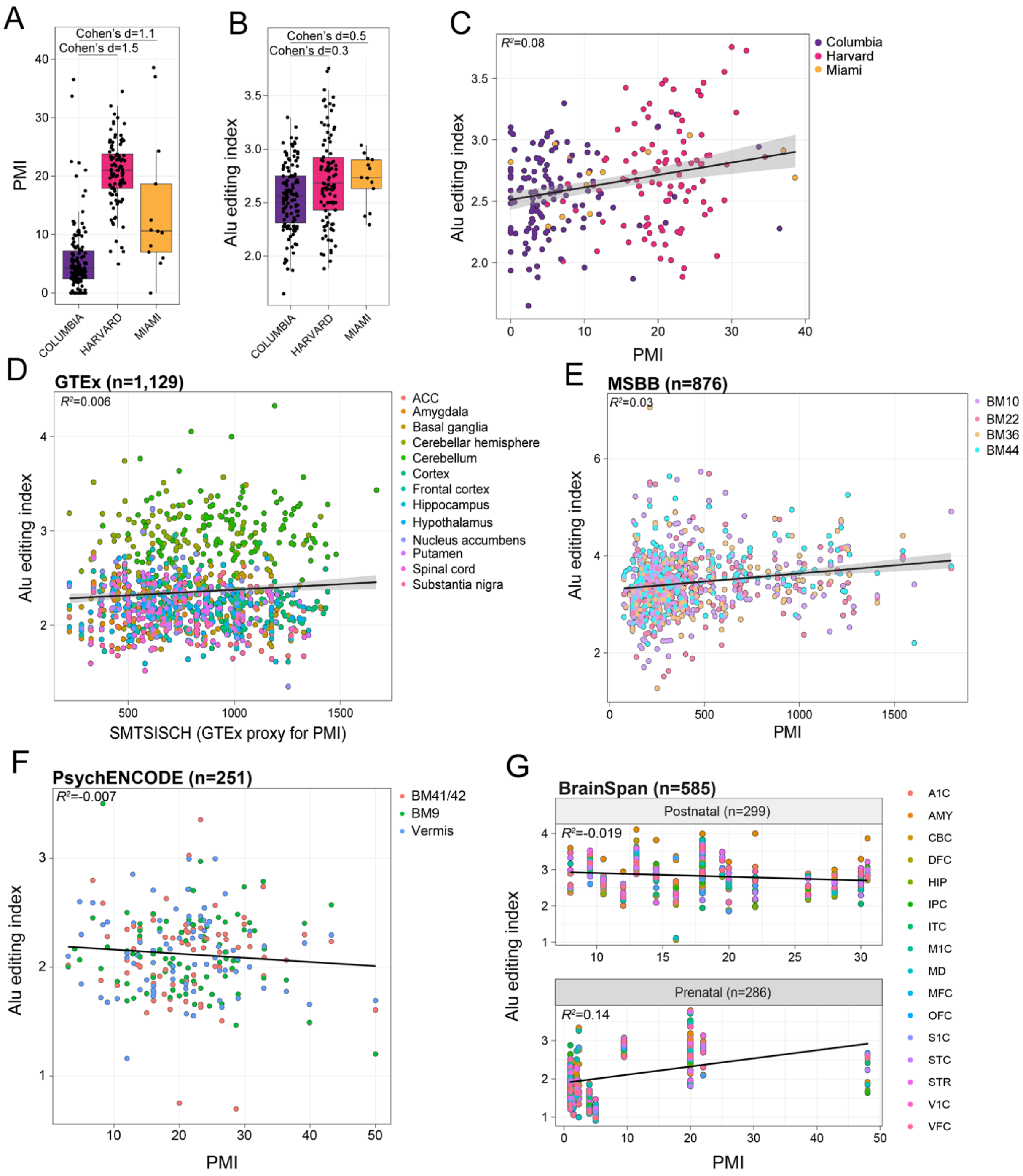


**Supplemental Figure 3. The relationship between extended postmortem interval and *Alu* editing.** (**A**) Differences in postmortem interval (PMI; y-axis) within three postmortem brain banks (x-axis) used in the Living Brain Project. (**B**) Differences in Alu editing index (AEI; y-axis) between the three postmortem brain banks (x-axis). Cohen's D was used to measure effect size differences between brain banks. All boxplots show the medians (horizontal lines), upper and lower quartiles (inner box edges), and 1.5× the interquartile range (whiskers). (**C**) Pearson's correlation coefficient of the relationship between PMI (x-axis) and the AEI (y-axis) in the Living Brain Project. (**D**) The relationship between PMI (x-axis) and the AEI (y-axis) for 1,129 bulk tissue postmortem RNA-seq samples across 13 brain regions. (**E**) The relationship between PMI (x-axis) and the AEI (y-axis) for 876 postmortem bulk RNA-seq samples across four cortical areas from the Mount Sinai Brain Bank (MSBB). A Pearson's correlation coefficient was used to test each association.

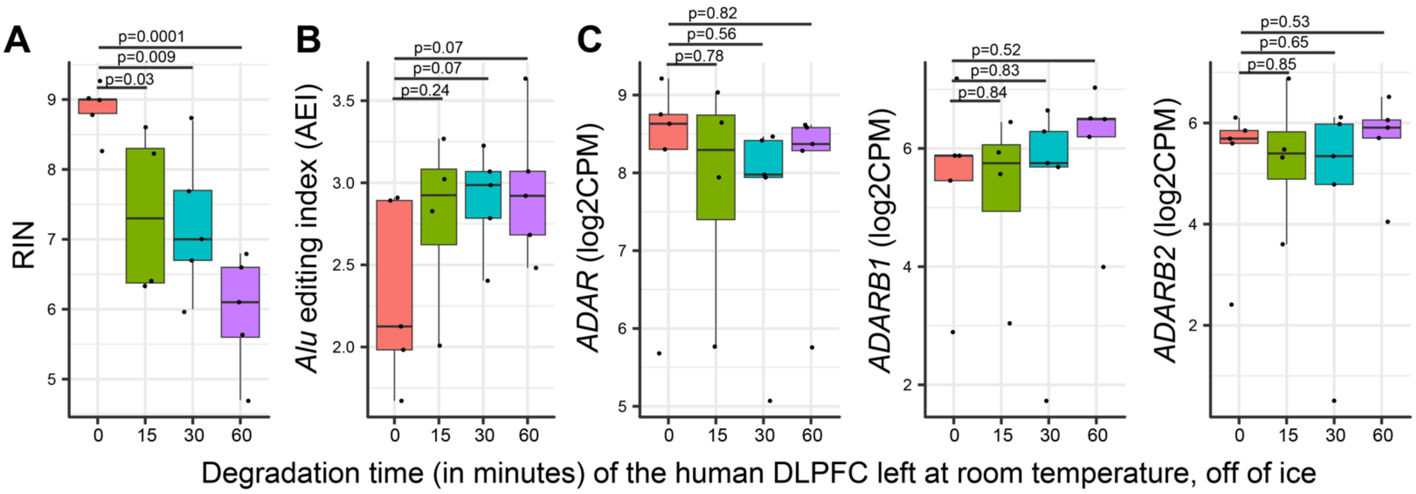


**Supplemental Figure 4**. **Quantification of RNA editing metrics throughout molecular degradation of the human DLPFC**. We downloaded existing RNA-sequencing data of a molecular degradation assay of the human DLPFC (PMID: 28634288). These data were prepared using RiboZero RNA-seq sequencing library, which align with the RNA-seq library preparation methods used in the current study. Quantification of (**A**) RNA integrity numbers (RINs) and (**B**) the AEI, as well as (**C**) *ADAR*, *ADARB1*, and *ADARB2* expression throughout advancing degradation of the DLPFC. Degradation of the DLPFC was measured in minutes left at room temperature (off of ice). For each measurement, a two-sided *t*-test compared 0 minutes (baseline) to each of the subsequent time-points without adjusting for multiple comparisons.

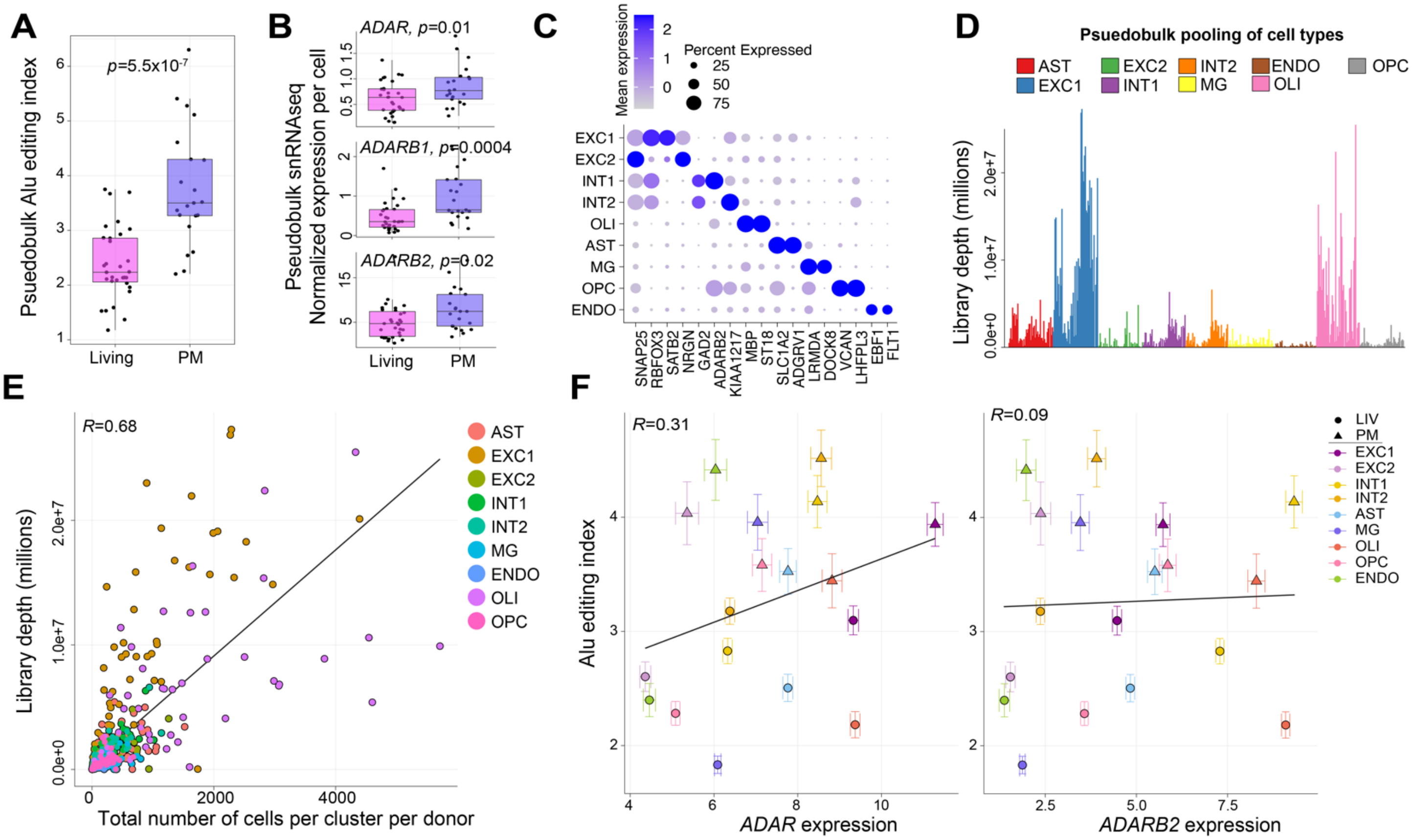

**Supplemental Figure 5. Pseudo-bulk snRNA-seq analyses.** (**A**) Leveraging snRNA-seq as pseudo-bulk tissue to measure the *Alu* editing index (AEI) between living and postmortem (PM) DLPFC. Two-sided linear regression was used to test for significance. (**B**) Raw counts of *ADAR*, *ADARB1* and *ADARB2* normalized to the total number of cells sequenced per donor, illustrating increased expression per cell across each gene in PM relative to living DLPFC. Mann-Whitney U test was used to test for significance. All boxplots in this figure show the medians (horizontal lines), upper and lower quartiles (inner box edges), and 1.5× the interquartile range (whiskers). (**C**) Dot plot of 17 cell marker genes used to define cell identities for nine cell populations identified via UMAP from snRNA-seq data. (**D**) Cell-type specific bam files were constructed for nine different cell types for each donor. Large differences in final library depth (y-axis) were observed across each pseudo-bulk cellular pool (x-axis). (**E**) Library depth for each pseudo-bulk cellular pool (y-axis) is correlated with the total number of cells per pseudo-bulk cellular pool per donor (x-axis). (**F**) Mean AEI (y-axes) relative to mean expression of *ADAR* (left) and *ADARB2* (right) for each pseudo-bulk cellular pool within living and PM DLPFC (x-axes). A Pearson's correlation coefficient was used to test each association. Standard error bars capture group-wise variance within living and postmortem tissues, respectively.

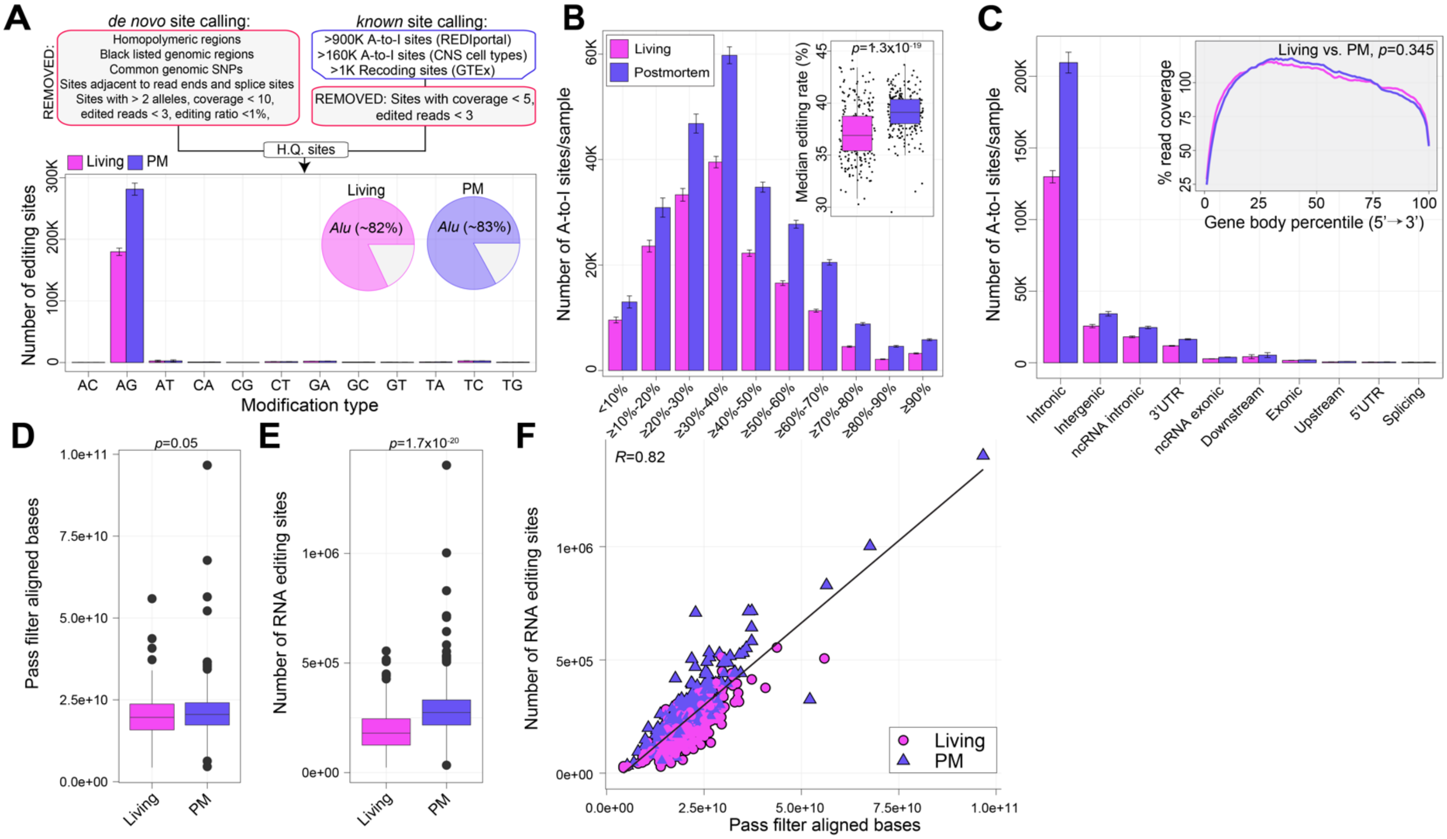


**Supplemental Figure 6. RNA editing site detection from bulk RNA-sequencing data.** (**A**) Uncovering high-quality (HQ) sites (top) using a combined de novo and supervised RNA editing site detection approach. Bar plots depict mean (with standard error) number of HQ sites for living (n = 164) and postmortem (PM) (n = 233) DLPFC samples based on substitution type and *Alu* repeat element (bottom). (**B**) The number of A-to-I sites (y-axis) binned by different editing levels in living and PM DLPFC (x-axis). Standard error bars show variance. Inset boxplots depict the median editing rates across all A-to-I sites per donor. Two-sided linear regression was used to test for significance. (**C**) Number of A-to-I sites detected per sample for living and postmortem tissues. Standard error bars show variance. Inset figure shows percentage of RNA-seq read coverage across each gene body (5' to 3'). Mann-Whitney U test was used to test for significance. (**D**) Median number of pass filter aligned bases (via Picard tools) (y-axis) and (**E**) median number of RNA editing sites per sample (y-axis) by living and PM DLPFC (x-axes). Two-sided linear regression was used to test for significance. All boxplots in this figure show the medians (horizontal lines), upper and lower quartiles (inner box edges), and 1.5× the interquartile range (whiskers). (**F**) Association between the number of RNA editing sites detected (y-axis) and the number of pass filter aligned bases (x-axis). A Pearson's correlation coefficient was used to test the association.

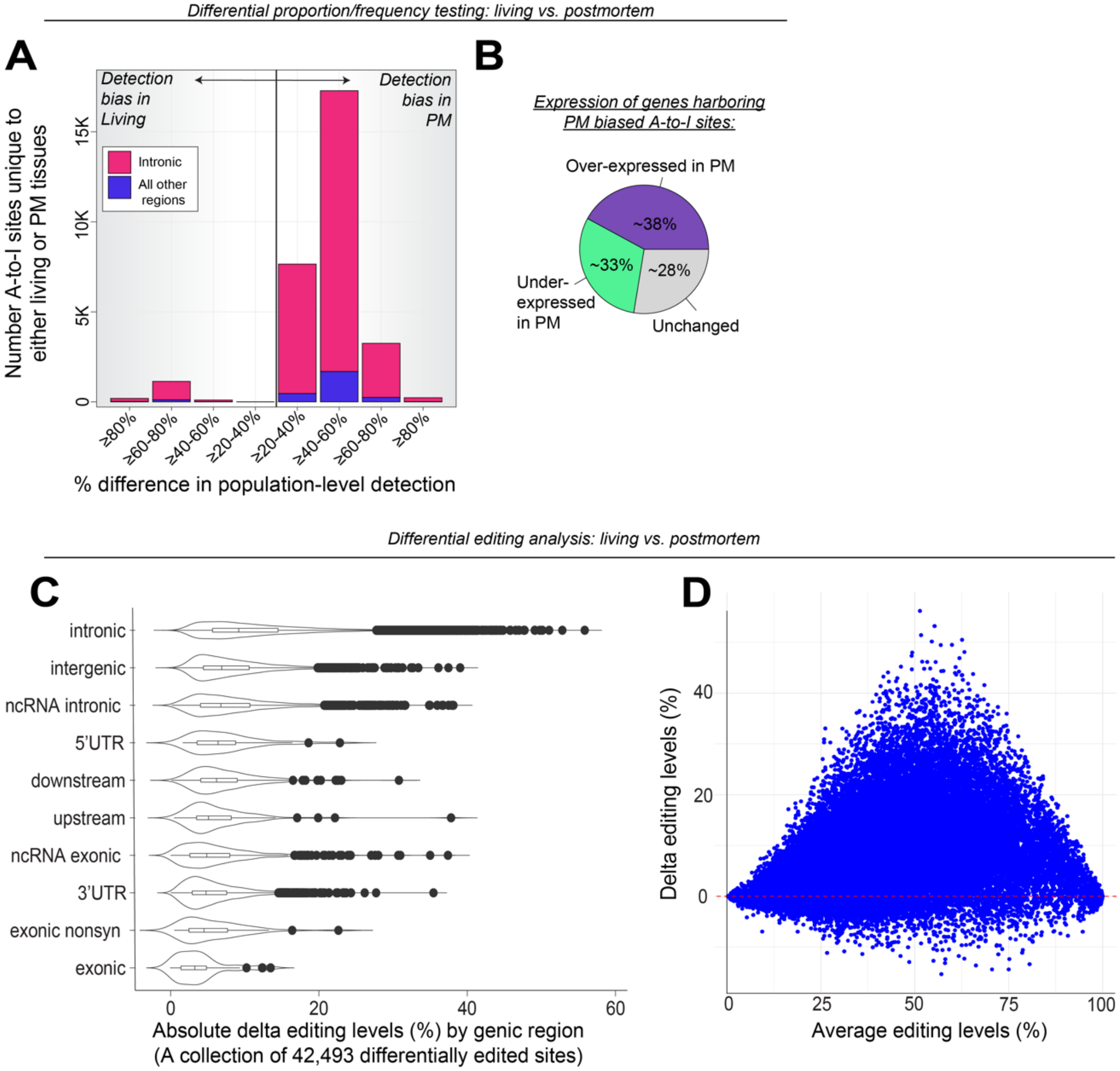


**Supplemental Figure 7. Differential testing between living and postmortem tissues.** Differential proportion testing: (**A**) The total number of A-to-I sites that are unique to either living or postmortem (PM) DLPFC (y-axis) according to their population-level detection (x-axis). For example, more than 16,824 A-to-I sites are detected in at least 40-60% of PM DLPFC and are not detected in living DLPFC. Most of these sites are intronic. (**B**) The expression patterns of genes harboring sites that are uniquely detected in PM DLPFC, indicate that overexpression of gene expression in PM tissues does not explain this enrichment. Differential editing analysis: (**C**) A collection of 42,493 differentially edited sites (FDR <5%) between living and postmortem DLPFC were used to plot the absolute delta values according to each genic region. All boxplots in this figure show the medians (horizontal lines), upper and lower quartiles (inner box edges), and 1.5× the interquartile range (whiskers). (**D**) MA-plot displaying delta editing levels (y-axis) against average editing levels (%; x-axis) across each transcriptomic sample. The red dotted line indicates 0% change in editing levels between living and postmortem tissues.

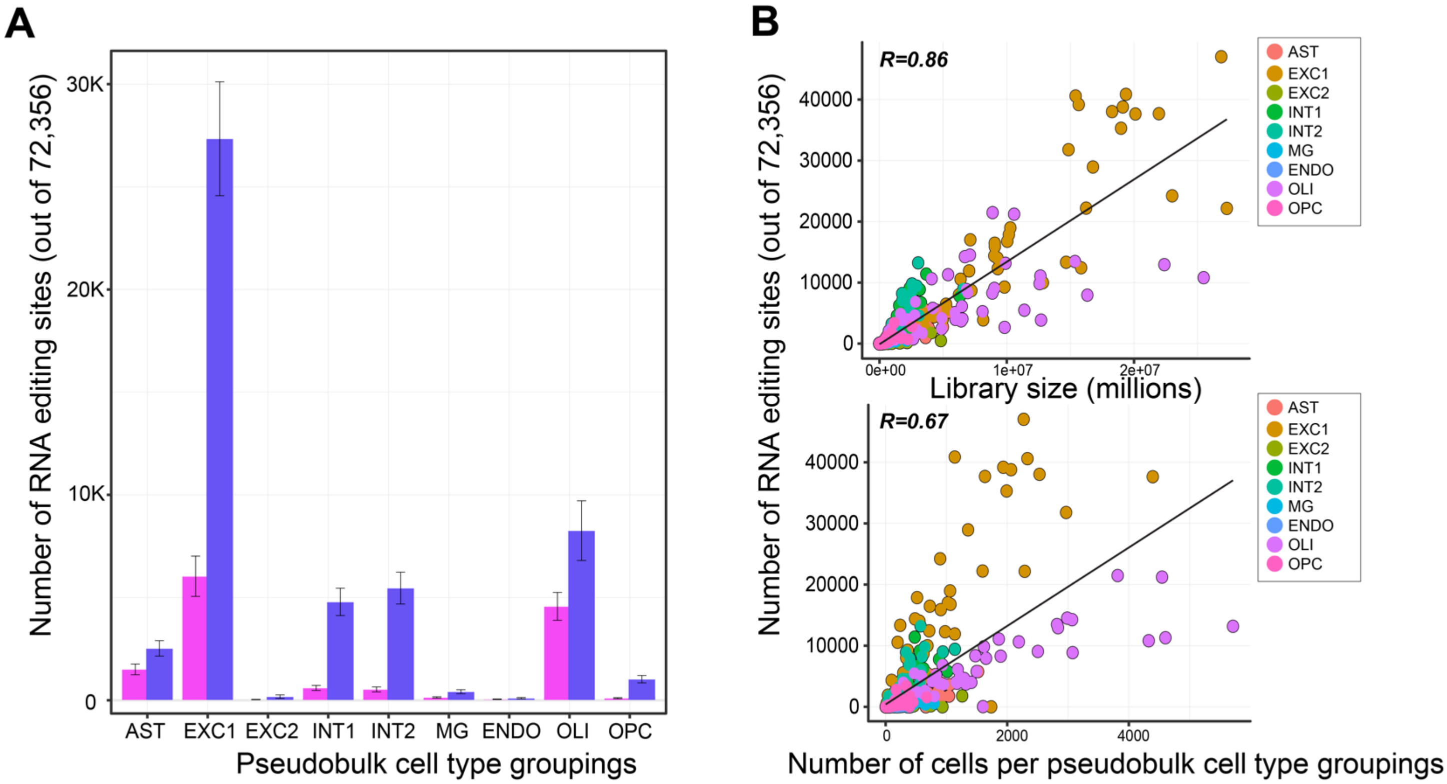


**Supplemental Figure 8. Querying LIV-PM sites in snRNA-seq cellular pools.** (**A**) We queried 72,356 A-to-I sites that were either differentially edited or differentially detected in either living or postmortem bulk RNA-seq DLPFC (y-axis) in snRNA-seq pseudo-bulk cellular pools (x-axis). The mean number of detected sites (with standard error bars) per pseudo-bulk cellular pool for each living and postmortem DLPFC are displayed. (**B**) The total number of A-to-I sites detected (out of 72,356) per pseudo-bulk cellular pool for sample (y-axes) as a function of library size (top, x-axis) and number of cells per pseudo-bulk cellular pool (x-axis, bottom). A Pearson's correlation coefficient was used to test the association.

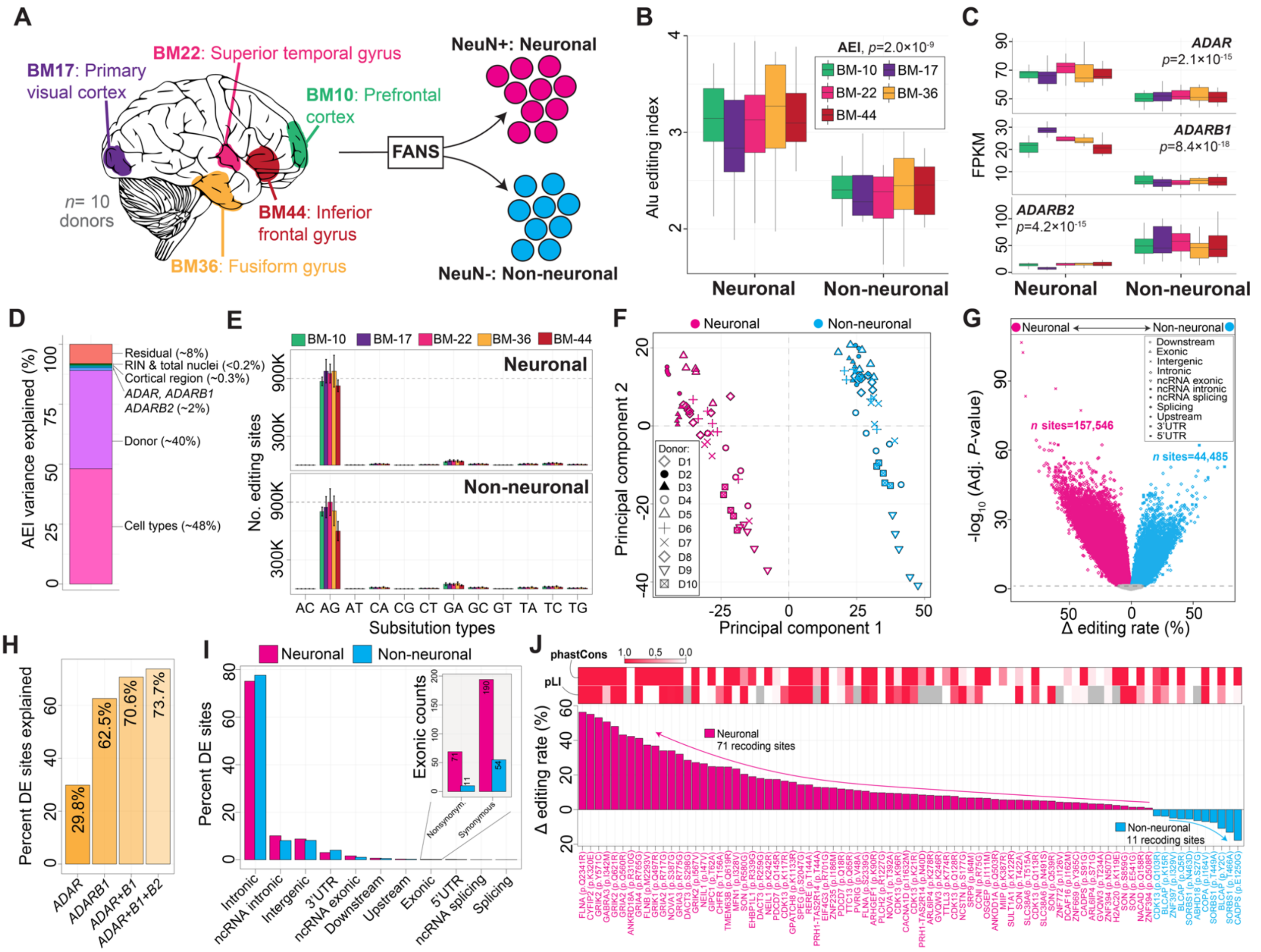


**Supplemental Figure 9. Neuronal and non-neuronal A-to-I editing across five postmortem cortical areas.** **(A)** Experimental design to generate deeply sequenced neuronal and non-neuronal nuclei across five cortical areas from ten biological replicates. (**B**) The *Alu* editing index (AEI) and (**C**) *ADAR*, *ADARB1* and *ADARB2* normalized expression within each cortical region between neuronal and non-neuronal nuclei. Two-sided linear regression was used to test for significance. Significance was tested between neuronal and non-neuronal cell types. No regional differences were observed. All boxplots in this figure show the medians (horizontal lines), upper and lower quartiles (inner box edges), and 1.5× the interquartile range (whiskers). (**D**) A linear mixed model quantified the fraction of AEI variance explained by eight known factors. (**E**) Mean number of RNA editing sites detected per sample according to different substitution types for neuronal (top) and non-neuronal nuclei (bottom). Standard error bars depict sample-level variation. (**F**) Principal component analysis of editing levels for 290,495 sites detected across neuronal and non-neuronal nuclei from all regions separates by cell type (PC1, x-axis) and donor as a repeated measure (PC2, y-axis). (**G**) Differential editing analysis compares the delta editing rates (%; x-axis) and the strength of significance (-log$_{10}$ adjusted p-value; y-axis) between neuronal and non-neuronal cell types. Neuronal biased sites are pink and non-neuronal sites are blue, and shapes represent unique genic regions. (**H**) Percentage of differential edited sites explained (y-axis) after covarying for different combination of ADAR enzymes as continuous measures (x-axis). (**I**) Percentage of differentially edited sites (y-axis) that map to different genic regions (x-axis). (**J**) Recoding sites that are either neuronal or non-neuronally biased ranked by their delta editing rate (%; y-axis) between cell populations. phastCons for each site and pLI for each gene were calculated and colored on a scale from 0-1.

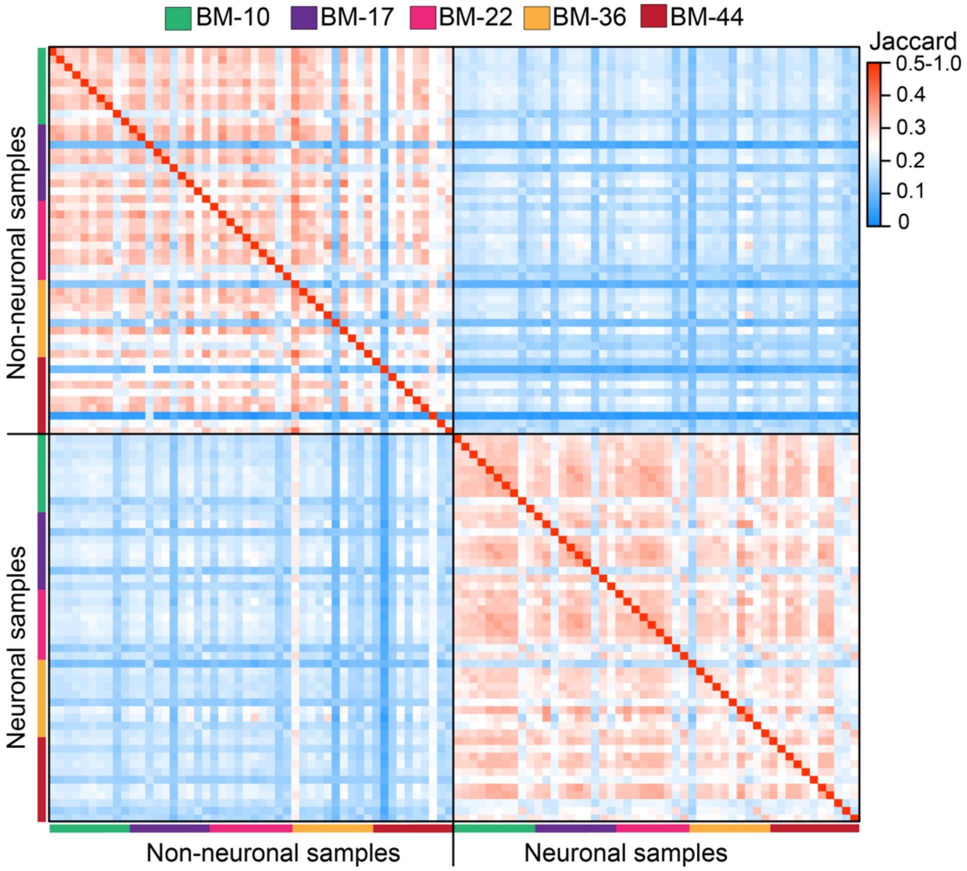


**Supplemental Figure 10. Jaccard overlap matrix of A-to-I sites per donor.** Pairwise overlaps for A-to-I sites detected for neuronal (NeuN+) and non-neuronal (NeuN-) cell populations across five cortical regions (n=10 biological replicates). Overlaps were measured using Jaccard Index and site detection converged mainly within either neuronal or non-neuronal nuclei, with little to no regional effect.

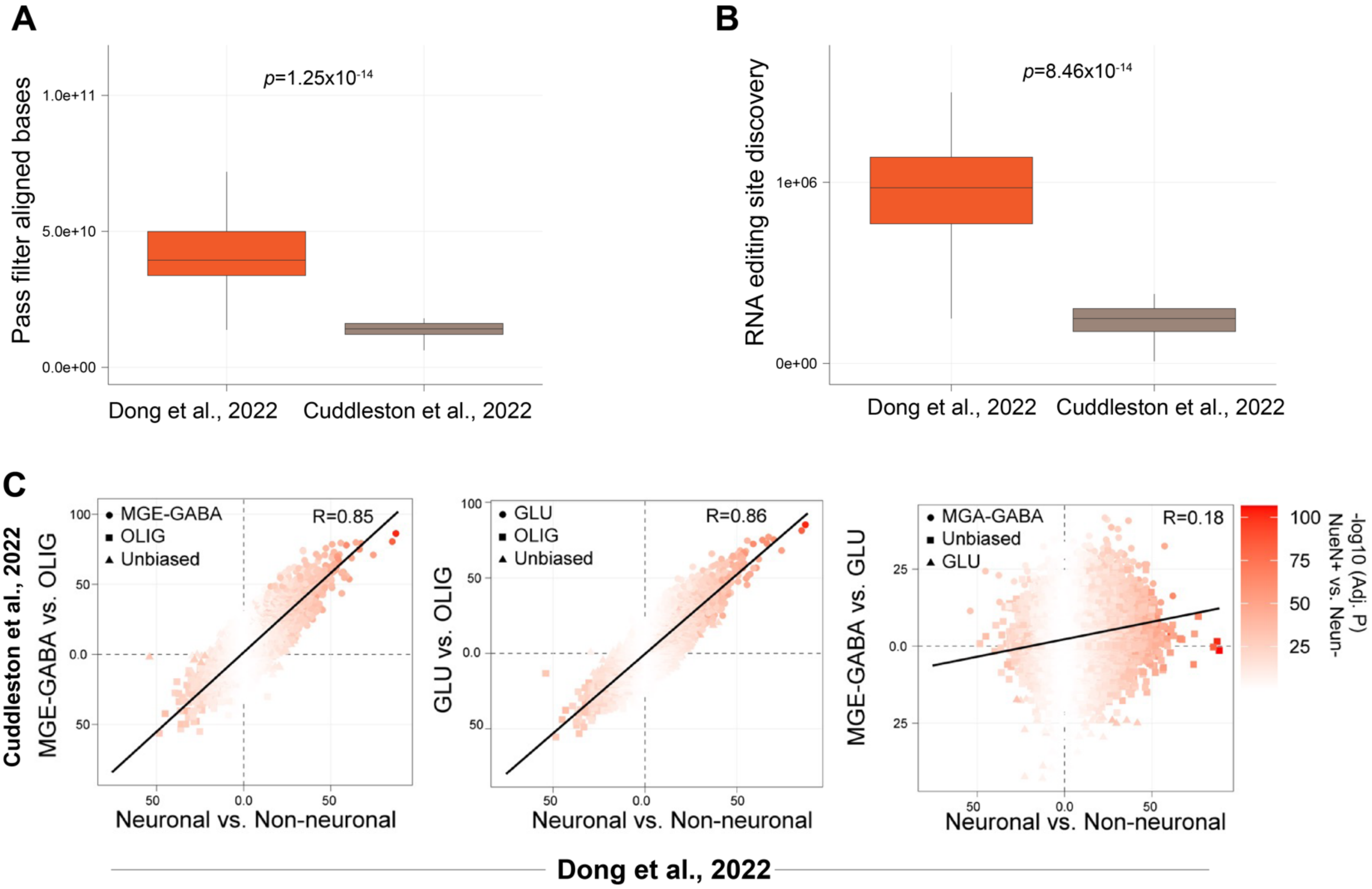


**Supplemental Figure 11. Validating cell-specific A-to-I sites.** To validate the specificity of our findings, we leveraged an independent RNA-seq study that performed fluorescence activated nuclei sorting (FANS) of MGE-GABAergic neurons (MGE-GABA), glutamatergic neurons (GLU) and oligodendrocytes (OLIG) from the human prefrontal cortex (*n*=9 biological replicates) (PMID: 35637184, Cuddleston et al., 2022). The FANS data in the current data set exhibits (**A**) significantly deeper sequencing depth and (**B**) significantly more A-to-I site discovery. Significance was tested with a Mann Whitney U test. (**C**) A total of 48,921 cell-specific sites were commonly detected between our data set and this previous study. Each site illustrated strong hallmark signatures to be either neuronal or non-neuronal and validated across studies. The neuronal vs. non-neuronal effect was compared with the MGE-GABA vs. OLIG effect (left), the GLU vs. OLIG effect (middle) and the MGE-GABA vs. GLU effect (right). All boxplots in this figure show the medians (horizontal lines), upper and lower quartiles (inner box edges), and 1.5× the interquartile range (whiskers).

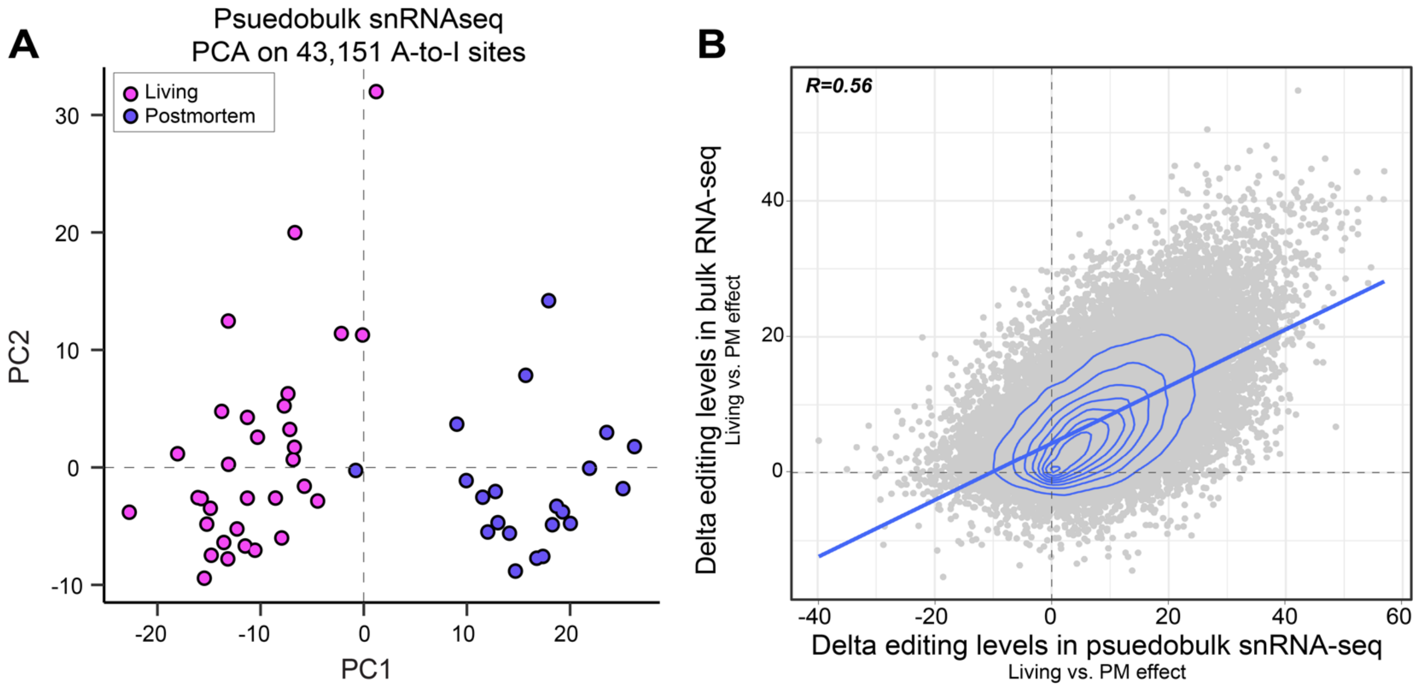


**Supplemental Figure 12. Replicating differentially edited sites using pseudo-bulk snRNA-seq. (A)** A total of 54,826 A-to-I sites detected across all bulk tissue samples were queried in pseudo-bulk snRNA-seq in an independent cohort of 21 living and 22 postmortem DLPFC samples. Of these, we detected 43,151 of these sites (), which were subjected to downstream analysis. (**A**) Principal component analysis (PCA) clearly separated living and postmortem DLPFC based on editing levels for these 43,151 sites along PC1. (**B**) Differential editing analysis was conducted (in an identical fashion as applied to bulk RNA-seq) to the pseudo-bulk profiles. To determine the degree of replication, the delta editing values obtained between living and postmortem DLPFC (x-axis) were regressed onto those derived from the bulk RNA-seq comparison (y-axis). This analysis demonstrated a positive concordance between the postmortem-induced effects of editing on these sites across the two data types and cohorts.

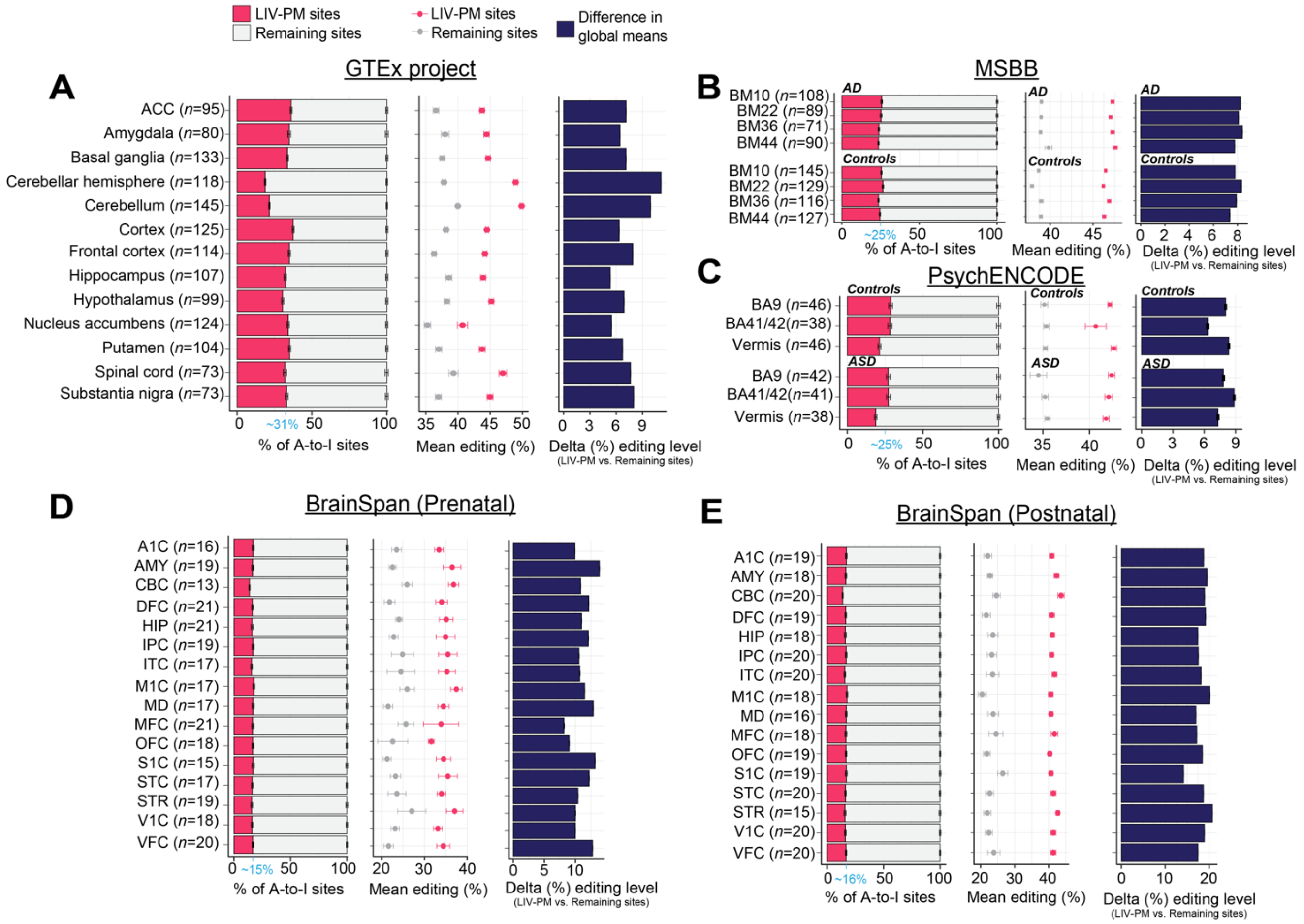


**Supplemental Figure 13. Quantifying postmortem effects in independent postmortem brain transcriptome data sets.** Four datasets were quality controlled and known A-to-I sites were quantified using a supervised approach requiring at least 5 reads and 2 edited reads (*see Materials and Methods*). For each dataset, we queried the fraction of sites catalogued as LIV-PM per sample (left), the global mean editing levels (%) for the LIV-PM sites relative to the remaining sites detected (middle) and the difference in global mean editing levels (LIV-PM site editing levels subtracted from the editing levels from the remaining sites (right)). These analyses were performed on **(A)** 13 anatomical regions from GTEx, (**B**) 4 anatomical regions from the Mount Sinai Brain Bank (MSBB), (**C**) 3 anatomical regions from PsychENCODE, and 16 anatomical regions from BrainSpan, covering (**D**) prenatal and (**E**) postnatal stages of development. MSBB samples included individuals with a clinical diagnosis of Alzheimer's disease (AD; CERAD scores indicative of definitely AD). PsychENCODE samples included individuals with a clinical diagnosis of autism spectrum disorder (ASD).

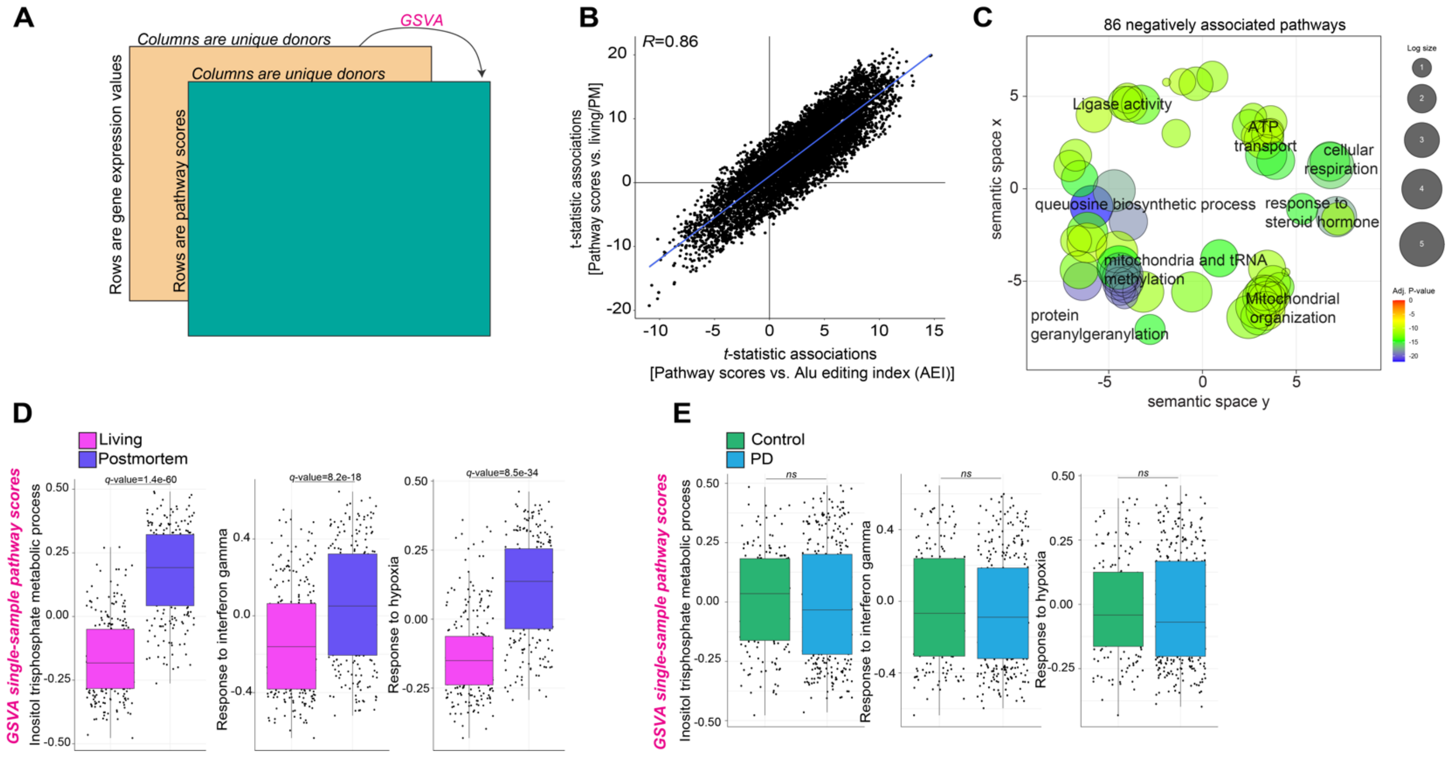


**Supplemental Figure 14. Single-sample gene-set variation analysis. (A)** The GSVA framework provides a summary measure of gene expression profiles underlying discrete biological processes by transforming an input gene-by-sample expression data matrix into a corresponding gene-set-by-sample expression data matrix. A total of 10,493 biological processes were generated for each living and postmortem bulk RNA-seq sample using the GSVA method. (**B**) Pathway activation scores were regressed onto the *Alu* editing index (AEI; x-axis) as well as differences between living and postmortem DLPFC (y-axis). (**C**) REVIGO semantic similarity analysis of the 86 pathways that negatively predict AEI. Examples of GSVA single-sample pathway scores (y-axes) that stratify (**D**) living versus postmortem tissues (x-axes) and (**E**) controls versus individuals with Parkinson's disease (PD). *Q*-values are multiple test corrected measures of significance (*see* ***Supplemental Data 2*** *for full summary statistics*).

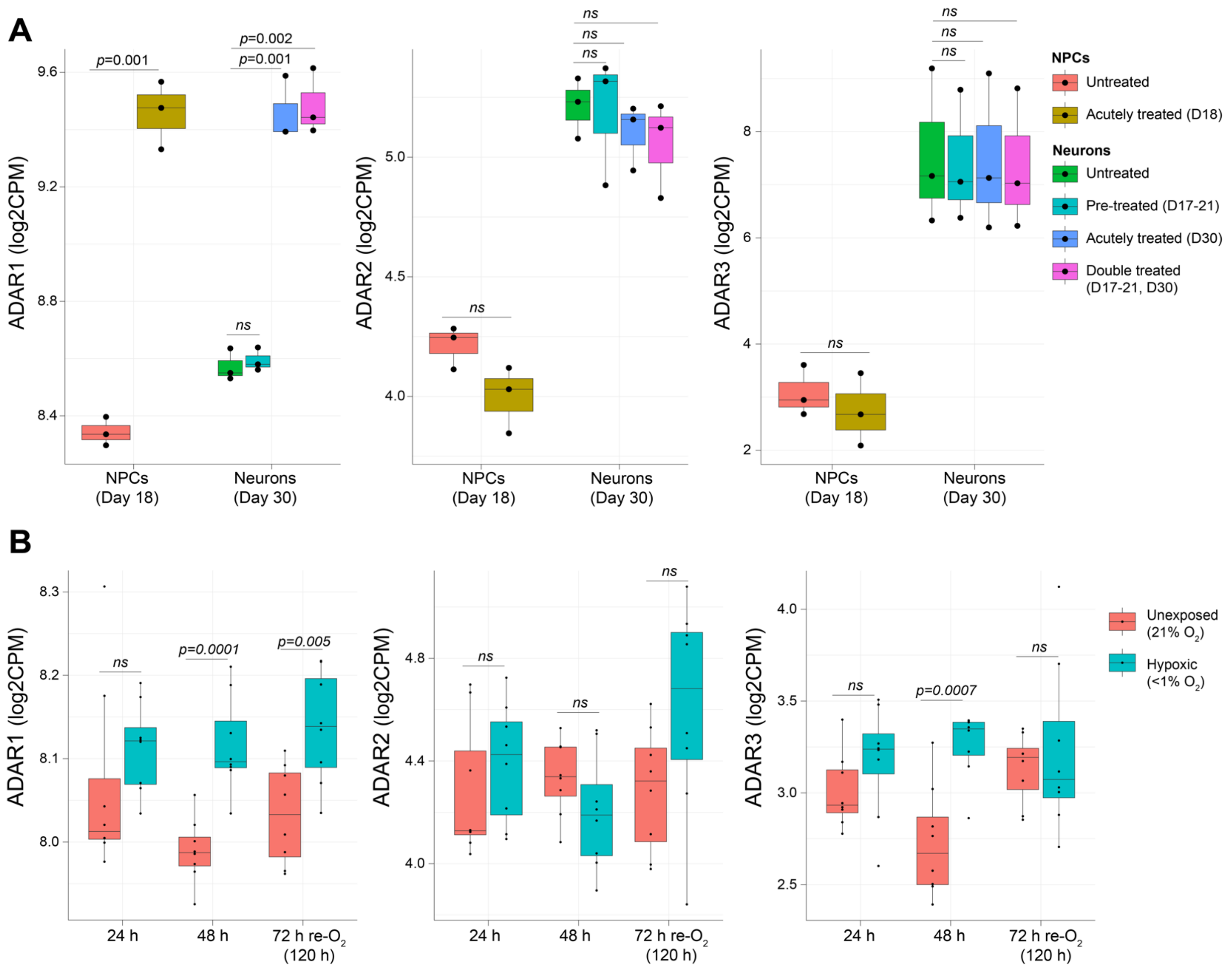


**Supplemental Figure 15. ADAR mRNA expression profiles.** VOOM normalized expression data were used to plot normalized expression of *ADAR1* (left), *ADAR2* (center) and *ADAR3* (right) across two in vitro models: **(A)** an in vitro model of hiPSC-derived neural progenitor cells (NPCs) and mature neurons treated with interferon-gamma; and (**B**) human cortical spheroids (hCS) exposed to hypoxic conditions (<1% $O_2$) and after 72 hours of reoxygenation versus unexposed (21% $O_2$). A two-sided student's *t*-test was used to test for significance. All boxplots in this figure show the medians (horizontal lines), upper and lower quartiles (inner box edges), and 1.5× the interquartile range (whiskers).

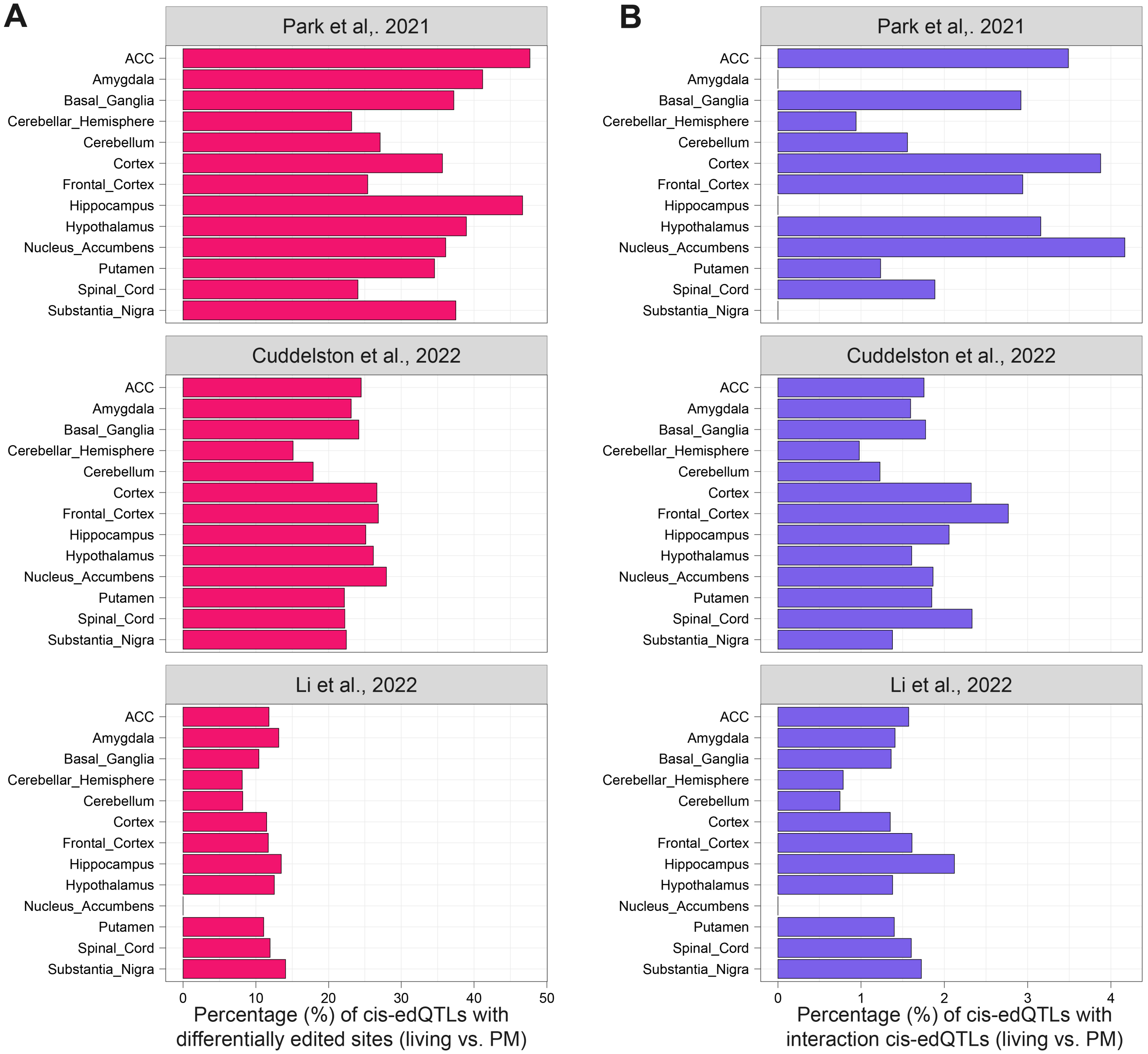


**Supplemental Figure 15. Annotation of independent cis-edQTLs from GTEX.** Three independent studies have quantified cis-edQTLs across 13 different GTEx brain regions. Here we show the percentage of each previously described set of eSites (*i.e.* A-to-I sites with at least one FDR significant cis-edQTL) that are either (**A**) differentially edited between living and postmortem DLPFC or (**B**) cis-edQTLs with a significant interaction effect between living and postmortem DLPFC. Ultimately, anywhere between 16-50% of previously described cis-edQTLs are associated with a differentially edited site between living and postmortem DLPFC and ~2% of all cis-edQTLs show interaction effects.
