## Supplemental Note for "Divergent landscapes of A-to-I editing in postmortem and living human brain"

**A deep atlas of neuronal and non-neuronal A-to-I editing sites across five postmortem cortical areas**

While global *Alu* editing rates can be accurately measured via snRNA-seq[1], it is challenging to ascertain individual cell-specific sites with high confidence due to individual differences in cell type composition and low sequencing depth, with reads covering only a fraction of the entire transcriptome[1,2] (**Figure S8**). A-to-I editing levels are known to be elevated in GABAergic and glutamatergic neurons relative to oligodendrocytes, and these differences are mostly, but not fully, explained by increased expression of *ADAR* and *ADARB1*[1]. Similar profiles have been observed in mice and *Drosophila*[3,4], which further support our findings. These contrasting profiles between neurons and non-neurons prompted us to integrate a deeply sequenced resource of neuronal and non-neuronal nuclei isolated from ten biological replicates across five postmortem cortical regions[5] (**Figure S9A**). Here, the depth of read coverage and the major cell types and anatomical regions profiled, provide an optimal framework cataloguing cell-type specific A-to-I sites in the human cortex – with the caveat that these are derived from postmortem tissues. Fluorescence activated nuclei sorting (FANS) was applied to an antibody against the well-established neuronal marker NeuN (*i.e.* RNA-binding protein RBFOX3) was used to isolate neuronal from non-neuronal nuclei followed by bulk RNA-sequencing. The AEI was computed for each sample and confirmed higher levels of global *Alu* editing in neuronal versus non-neuronal nuclei ($p$=2.0×$10^{-9}$) (**Figure S9B**). No significant differences were observed in global *Alu* editing between cortical areas within each cellular population. The expression of *ADAR* and *ADARB1* was significantly higher in neuronal nuclei ($p$=8.4×$10^{-15}$, $p$=2.1×$10^{-18}$, respectively) and positively correlated with the AEI ($r$=0.47, $r$=0.52, respectively), while *ADARB2* was more highly expressed in non-neuronal nuclei ($p$=4.2×$10^{-15}$) and negatively correlated with the AEI ($r$=-0.51) (**Figure S9C**). A linear mixed model quantified the variance of the AEI explained by known factors, and cell type differences explained the largest median fraction of *Alu* editing variability (~48%), followed by donor as a repeated measure (~40%) (**Figure S9D**).

To quantify individual cell-specific sites, we applied the same pipeline as above to query high-confidence A-to-I sites across neuronal and non-neuronal nuclei (**Supplemental Data 4**). A mean of 1,036,579 editing sites were detected per sample across neuronal nuclei and 925,440 sites were detected per sample across non-neuronal nuclei (**Figure S9E**), which showed similar hallmarks of ADAR-mediated RNA editing, in that the majority: (1) were A-to-I sites (~84% neuronal, ~88% non-neuronal); (2) mapped to *Alu* repeats (~75% neuronal, ~75% non-neuronal); (3) were predominately known sites cataloged in editing databases (~78% neuronal, ~80% non-neuronal). The enrichment of A-to-I sites in non-coding regions is consistent with existing reports that intronic regions are over-represented in RNA-seq and snRNA-seq generated from the nuclear fraction relative to a combined cytoplasmic and nuclear fraction[6-8]. In general, site detection was convergent between neuronal and non-neuronal populations, with little-to-no regional specificity (**Figure S10**). PCA applied to the editing levels of 290,495 sites detected across all nuclei and regions confirmed a significant cell-type effect along PC1 and a donor effect along PC2 (**Figure S9F**). Differential editing tested for differences in mean editing levels between neuronal and non-neuronal nuclei: 157,546 sites displayed higher editing levels in neurons ('neuronal-biased') and 44,485 sites displayed higher editing levels in non-neuronal cells (non-neuronal biased) (**Figure S9G**). Covarying for *ADARB1* and *ADAR* expression explained a large fraction of differentially edited sites (**Figure S9H**). Most differentially edited sites mapped to non-coding regions (**Figure S9I**). A total of 82 recoding sites were differentially edited; 71 were neuronal-biased, which were commonly evolutionary conserved (phastCons) and mapped to genes with elevated pLI, underscoring their putative functionality (**Figure S9J**). Finally, we replicated these findings and significantly expand the scope of existing catalogues of cell-specific RNA editing in the cortex[1] (**Figure S11**), offering 201,941 A-to-I sites specific to either neuronal or non-neuronal nuclei. All results are presented in **Supplemental Data 4**.

### REFERENCES RELATED TO SUPPLEMENTAL NOTE 1
